# White Matter is Increased in the Brains of Adults with Neurofibromatosis 1

**DOI:** 10.1101/2021.06.13.21258853

**Authors:** Su Wang, Jan M. Friedman, Per Suppa, Ralph Buchert, Victor-Felix Mautner

## Abstract

**Objective:** To characterize alterations in brain volumes by three-dimensional (3D) MRI in adults with neurofibromatosis 1 (NF1).

**Methods:** We obtained brain volume measurements using 3D magnetic resonance imaging for 351 adults with NF1 and, as a comparison group, 43 adults with neurofibromatosis 2 (NF2) or Schwannomatosis. We assessed a subset of 19 adults with NF1 for clinical severity of NF1 features and neurological problems and conducted psychometric testing for attention deficiencies and intelligence quotient. We compared brain volumes between NF1 patients and controls and correlated volumetric measurements to clinical and psychometric features in the NF1 patients.

**Results:** Total brain volume and total and regional white matter volumes were all significantly increased in adults with NF1. Grey matter volume decreased faster with age in adults with NF1 than in controls. Greater total brain volume and white matter volume were correlated with lower attention deficits and higher intelligence quotients in adults with NF1.

**Interpretations:** Our findings are consistent with the hypothesis that dysregulation of brain myelin production is a cardinal manifestation of NF1 and that these white matter changes may be functionally important in affected adults.

## INTRODUCTION

Neurofibromatosis 1 (NF1) is an autosomal dominant disease that affects around 1:3000 live births^1^. The disease is characterized by a wide range of features including café-au-lait macules, neurofibromas, central nervous system gliomas, learning disabilities, and attention deficits^2, 3^.Dysregulated growth of myelin-producing Schwann cells due to *NF1* loss of function is a hallmark of the disease^4, 5^.

Several studies have examined changes in brain morphology in people with NF1, but most of these studies have small sample sizes and focus on affected children^6-22^. The most consistent findings in individuals with NF1 have been increased total brain volume, total white matter volume and corpus callosum (CC) area^7-22^. Some studies also found increased grey matter volume in children or young adults with NF1^7, 8, 17^. Other studies have examined grey matter and white matter composition of regional cranial structures, with the most consistent observation being increased corpus callosum (CC) area and size^9, 11, 15, 16, 18, 20-22^.

We previously examined brain morphology using planar MRI in a large group of adults with NF1 (n = 389)^22^. We found an increase in CC area as well as an increase in brainstem size in comparison to unaffected controls. These structures are mainly composed of white matter, supporting previous findings of white matter enlargement in individuals with NF1.

Individuals with NF1 have cognitive and behavioural abnormalities more frequently than expected. The average intelligence quotient (IQ) of people with NF1 is about one standard deviation lower than the average in general population^3^. Learning disability occurs in 30% to 70% in individuals with NF1 compared to 8% in the general population, and attention deficit hyperactivity disorder (ADHD) occurs in ∼ 40% in individuals with NF1 compared to ∼10% of the general population^23-26^.

Several studies have attempted to correlate changes in brain morphology and volume with cognitive or behavioural abnormalities in individuals with NF1. All of these studies, except one (small sample size using 2D brain morphology measurements)^22^, were conducted in affected children. As well, the correlations found were inconsistent across studies, perhaps due to limited sample sizes and numerous variables examined^7-10, 14, 15, 17, 19-21^. Our previous study attempted to correlate CC and brainstem size to the severity of NF1, neurological features, and ADHD, as well as IQ and attentional deficits, but found no associations^22^.

In the current study, we compared white matter and grey matter volumes in the whole brain and in major intracranial sub-regions in adults with NF1 and controls. We also assessed the effect of age on these volumes and correlated brain white matter and grey matter volumes with neuropsychometric findings in adults with NF1.

## METHODS

### Participants

Between 2003 and 2015, all patients seen in the NF Outpatient Department of the University Hospital Hamburg-Eppendorf in Hamburg, Germany were offered brain MRI to monitor their intracranial tumour burden. Three-dimensional (3D) magnetization-prepared rapid acquisition with gradient echo (MPRAGE) brain volumetric scans were obtained for 351 adults with NF1 and 43 adults with Neurofibromatosis 2 (NF2) or Schwannomatosis. Adults with NF2 or Schwannomatosis were used as a comparison group as neither condition affects brain volume^27^. Participants were excluded if their MRI scans for two or more of the 45 volumetric measurements were classified as an outlier. This was done to detect and exclude brain lesions and gliomas. An outlier scan is classified below. Age and sex were also recorded at the time of each examination.

A subset of the adults with NF1 (n = 19) also received formal ratings for NF1 severity, neurological severity, and ADHD severity, as well as neuropsychometric assessments for attention deficits and IQ.

### Brain Volumetric Measurements

#### MRI Preprocessing

MRI brain scans were preprocessed using MATLAB and the Statistical Parametric Mapping toolbox (version SPM12, Wellcome Trust Centre for Neuroimaging, London, UK) (www.fil.ion.ucl.ac.uk/spm). The unified segmentation engine of SPM12 was used to segment the MPRAGE MRI into grey matter (GM), white matter (WM) and cerebrospinal fluid (CSF). Default parameters for segmentation were used except that image data were sampled every 2 mm instead of the default 3 mm^28^. Diffeomorphic anatomical registration through exponentiated Lie algebra (DARTEL) was used for stereotactical normalization of the brain scans into the anatomical standard space of the Montreal Neurological Institute (MNI)^29^. GM and WM component images in native patient space obtained by unified segmentation were resliced to 0.5 mm voxel size prior to the DARTEL registration process in order to avoid aliasing artifacts^30^. DARTEL was used with default parameter settings and the IXI555 templates in MNI space provided by the VBM12 toolbox (http://dbm.neuro.uni-jena.de/vbm). Intensity modulation was applied to the normalized component images to preserve the overall volume.

#### MRI-based Volumetry

Volumetric measures were derived from the modulated tissue component images in MNI space. Total GM, total WM and total CSF volume were obtained by summing all voxel intensities in the corresponding component images and then multiplying the sum by the voxel volume. The total intracranial volume (TIV) was estimated by summing total GM, total WM and total CSF in a TIV mask predefined in MNI space^31^. Regional GM volume was determined for the following regions of interest (ROI): frontal lobe, parietal lobe, occipital lobe, temporal lobe, cerebellum, inferior frontal gyrus (IFG), insula, hippocampus without subiculum (HC), hippocampus with subiculum (HCS), amygdala, caudate, putamen, and thalamus. Regional WM volume was determined for the following ROIs: frontal lobe, parietal lobe, occipital lobe, temporal lobe, cerebellum, IFG, CC, brainstem, and pons. All ROIs were unilateral and were analysed separately in the left and in the right hemisphere except the ROIs for brainstem, pons, and CC.

The GM or WM volume in a given ROI was obtained by multiplying the corresponding modulated tissue component image (GM or WM) in MNI space with a binary mask of the ROI predefined in MNI space (voxel intensity = 1 for voxels belonging to the ROI, voxel intensity = 0 otherwise), summing all voxels intensities in the resulting masked image, and then multiplying the sum by the voxel volume.

The binary ROI masks were obtained from different brain atlases. The binary masks for frontal, parietal, occipital, temporal lobe, and the cerebellum were derived from the cerebral lobes atlas in MNI space^32^. The binary ROI masks for IFG, insula, caudate nucleus, putamen, and brainstem were derived from the LPBA40 atlas^33^. The ROI mask of the hippocampus without subiculum comprised cornus ammonis and fascia dentata^34^. The ROI mask of the hippocampus with subiculum included the subiculum in addition to cornus ammonis and fascia dentata^34^. The binary ROI masks for amygdala, thalamus and pons were derived from the Automated Anatomic Labeling atlas (amygdala, thalamus) and from the TDlobes atlas (pons), both implemented in the PickAtlas tool of Wake Forest University^35^. The binary CC mask was obtained as the union of binary masks for genu, body and splenium taken from the ICBM-DTI-81 white matter labels atlas^36, 37^. All masks were interpolated to 1.5 mm isotropic resolution to match the resolution of modulated and normalized component images.

The volumetric analyses were performed in batch mode, all MRI scans in a single batch, without visual quality control of spatial normalization and tissue segmentation. Thus, failure of spatial normalization and/or tissue segmentation could not be ruled out. Furthermore, brain lesions that might affect MRI-based volumetry could also not be ruled out. In order to detect and exclude these cases, an MRI scan was considered an ‘outlier’ with respect to a given volumetric measure if the value of this volumetric measure estimated from this scan was below (lower quartile – 2 x interquartile range) or above (upper quartile + 2 x interquartile range) over all MRI scans. This was tested separately for each volumetric measure. An MRI scan, including all volumetric measures derived from it, was excluded from all further analyses if it was an outlier with respect to two or more of the 45 volumetric measures. 125/863 MRI scans (14.5%) were identified as outlier and excluded. Some scans were repeat scans of the same participant.

### Clinical and Neuropsychometric Assessments

The clinical assessments [NF1 severity (n = 19), neurological severity (n = 19), ADHD diagnosis (n= 16)], and psychometric assessments [IQ (n = 16) and attention deficit [ACS] (n = 19)] were conducted in the same manner as previously described (Wang et al. 2021)^22^.

### Statistical Analysis

All analyses were conducted in R studio 3.6.0.

#### Demographic Analysis

Mean age was compared between adults with NF1 and the comparison group using the Student T test. Normality and equality of variance were established using the Shapiro-Wilk test and Fisher’s F test, respectively. Sex ratios were compared between adults with NF1 and the comparison group using the χ^2^ test.

#### Volumetric Analysis

Median brain volumes were compared between adults with NF1 and the comparison group using Mann-Whitney U tests. For some regions, volumes were not normally distributed or the average variance was not the same between hemispheres, so we used the more conservative non-parametric Mann-Whitney U test for all volume comparisons for consistency throughout. Significance values were adjusted using false discovery rate (FDR) to account for 38 volumetric comparisons.

#### Brain Volume to Age Regression Analysis

Multiple linear regression was used to determine the effect of sex, age, and NF1 status (as independent variables) on the total brain volume, GM volume, or WM volume, respectively (as the dependent variable). For total brain volume and WM volume, multiple linear regressions were separately conducted between individuals younger than 45 years old and older than 45 years old. Sex and NF1 status were coded as binary categorical data. The regression models were built using a forward stepwise method, adding sex, then age, then NF1 status sequentially. The models were compared using ANOVA.

#### Brain Volume to Neuropsychometric Correlation Analysis

Select brain volume measurements (total brain volume, GM, WM, white matter CC, white matter brainstem, left and right grey matter frontal lobe, left and right grey matter cerebellum, and left and right grey matter hippocampus without subiculum) were correlated to clinical severity and neuropsychometric measurements using Spearman or Pearson correlation (n = 16 to 19). All brain volume measurements were analysed as continuous data. Clinical assessments (NF1 severity, neurological severity, and ADHD diagnosis) were analysed as ordinal data. Neuropsychometric assessments (IQ and attention deficit [ACS]) were analysed as continuous data. Spearman’s Rank-Order correlation was conducted for ordinal data and Pearson correlation for continuous data. Significance values were adjusted using FDR to account for all correlation comparisons (n = 16 to 19).

### Data Availability

Anonymous data are available for appropriate research purposes through V.F. Mautner, MD.

## RESULTS

### Age and Sex Demographics of Study Participants

Both age and sex have been shown to affect brain volume in adults^38, 39^. Thus, it is important to determine whether there is a difference in the demographic composition between the NF1 and the comparison group to avoid the effect of sex and age as confounding factors in the analysis. This study included 351 adult participants with NF1 and 43 adult participants with either NF2 or Schwannomatosis (comparison group). Neither age distribution nor sex ratio was different between the two groups (Table 1).

**Table 1:** Participant demographics for adults with NF1 and comparison group (NF2 orSchwannomatosis).

|  | ADULTS WITH NF1 | COMPARISON GROUP | DIFFERENCE |
| --- | --- | --- | --- |
| N | 351 | 43 |  |
| Mean Age (SD) | 37.5 (13.2) | 40.0 (14.4) | Not significant |
| Age Range | 18.0 – 79.2 | 20.1 – 66.4 |  |
| Female: Male<br>(Female Percentage) | 205: 146<br>(58%) | 21:22<br>(48%) | Not significant |

### White Matter Brain Volume Is Increased in Adults with NF1

NF1 is characterized by the dysregulation of myelin formation^4, 5^. In the central nervous system, oligodendrocytes create an insulating myelin sheath around axons^40^. In the brain, myelination is mainly in white matter. To examine the effect of NF1 on myelination in the brain, we investigated the differences in brain volumes (total and regional white and grey matter) between adults with NF1 and individuals without NF1.

Adults with NF1 have significantly greater total brain volume and total white matter volume compared to the comparison group (Figure 1). As well, all the white matter regional measurements (frontal lobe, parietal lobe, IFG, CC, pons, and brainstem) showed a significantly increased volume in adults with NF1 compared to the comparison group (Figure 2). This increase in white matter volume was consistent bilaterally. We did not find a difference in total grey matter volume between the NF1 and comparison groups (Figure 1). However, we found some region-specific changes in grey matter volume. Seven of the 13 regions we measured were significantly larger in adults with NF1 than in the comparison group (Figure 3). These grey matter volume increases were also consistent bilaterally in all regions. These results demonstrate that adults with NF1 have uniformly increased white matter volume, and thus more myelin formation, than expected. NF1 also affects grey matter volume, but only in some of the smaller regions of the brain, so that total grey matter volume was not significantly different between adults with NF1 and controls.

**Figure 1:**
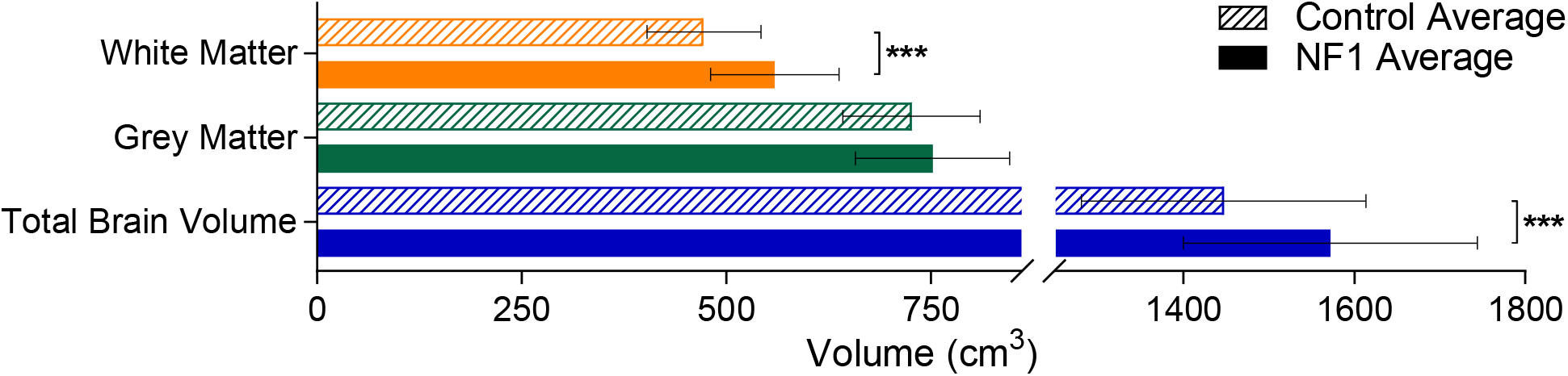
Average total brain volume, grey matter volume, and white matter volume for adults with NF1 and comparison group. Colored bars and outlines indicate different regions measured. Bar fill indicates participant group. Error bars indicate one standard deviation, and asterisks indicate FDR-adjusted statistical significance. *** = p < 0.001.

**Figure 2:**
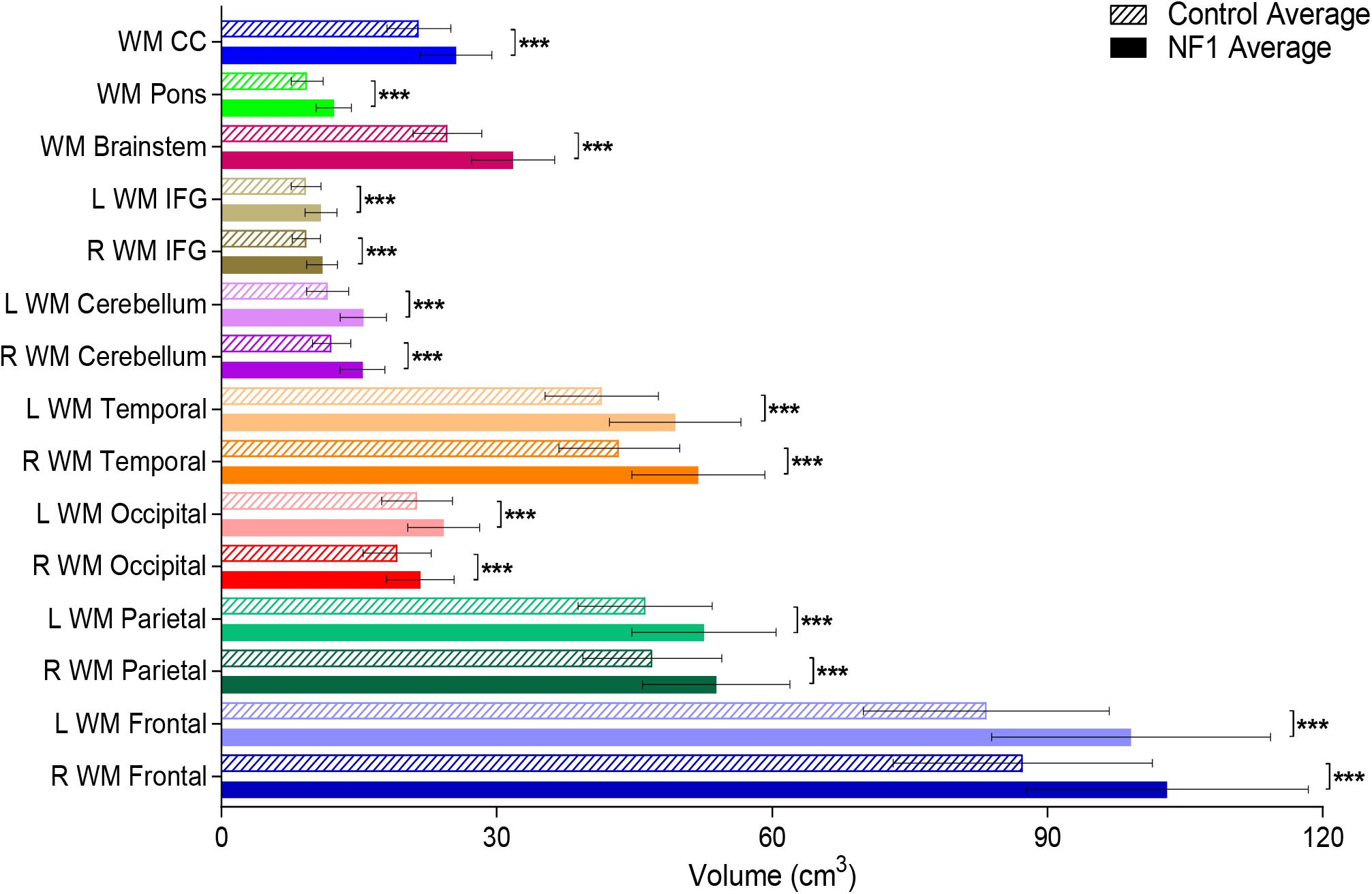
Average regional white matter brain volume for adults with NF1 and comparison group. Colored bars and outlines indicate different regions measured. Bar fill indicates participant group. Error bars indicate one standard deviation, and asterisks indicate FDR-adjusted statistical significance. *** = p<0.001. WM = white matter, R = right, L = left, CC = corpus callosum, IFG = inferior frontal gyrus.

**Figure 3:**
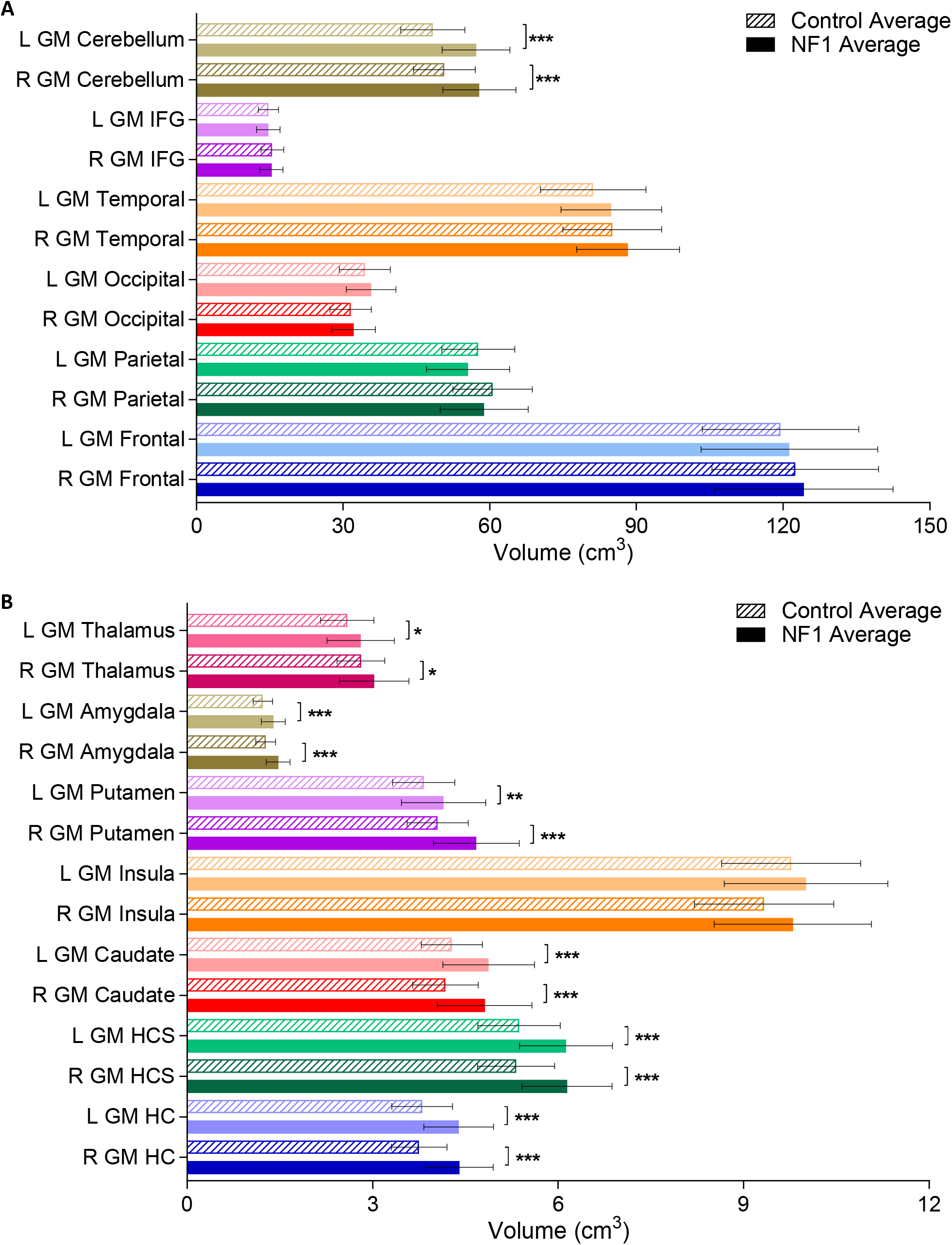
Average major (A) and minor (B) regional grey matter brain volume for adults with NF1 and comparison group. Colored bars and outlines indicate different regions measured. Bar fill style indicates participant group. Error bars indicate one standard deviation, and asterisks indicate FDR-adjusted statistical significance. * = p < 0.05, ** = p < 0.01, *** = p < 0.001. GM = grey matter, R = right, L = left, IFG= inferior frontal gyrus, HC = hippocampus without subiculum, HCS = hippocampus with subiculum.

### Grey Matter Volume Decreases Faster with Age in Adults with NF1 than Comparison Group

In the general population, grey matter volume decreases linearly over age. Meanwhile, total brain volume and white matter volume increases over age from early adulthood to around 45 years old, then decreases afterwards^38^. We were interested in whether the rate of reduction in total brain volume, grey matter, and white matter volume over age is different in adults with NF1 than in the general population. We regressed total brain volume, grey matter volume, and white matter volume, against age, separated by sex and NF status. We did not find an effect of age on either total brain volume or total white matter volume, either when regressed to the entire age spectrum or separated into younger than or older than 45 years old. (Figure 4A, B). We found that total grey matter volume decreased with age, with the adults with NF1 declining significantly faster than the comparison group (comparison group: β = -3.2; p = 0.00, between NF1 and comparison group: p = 0.03) (Figure 4C). These results demonstrate that people with NF1 have increased age related grey matter volume reduction.

**Figure 4:**
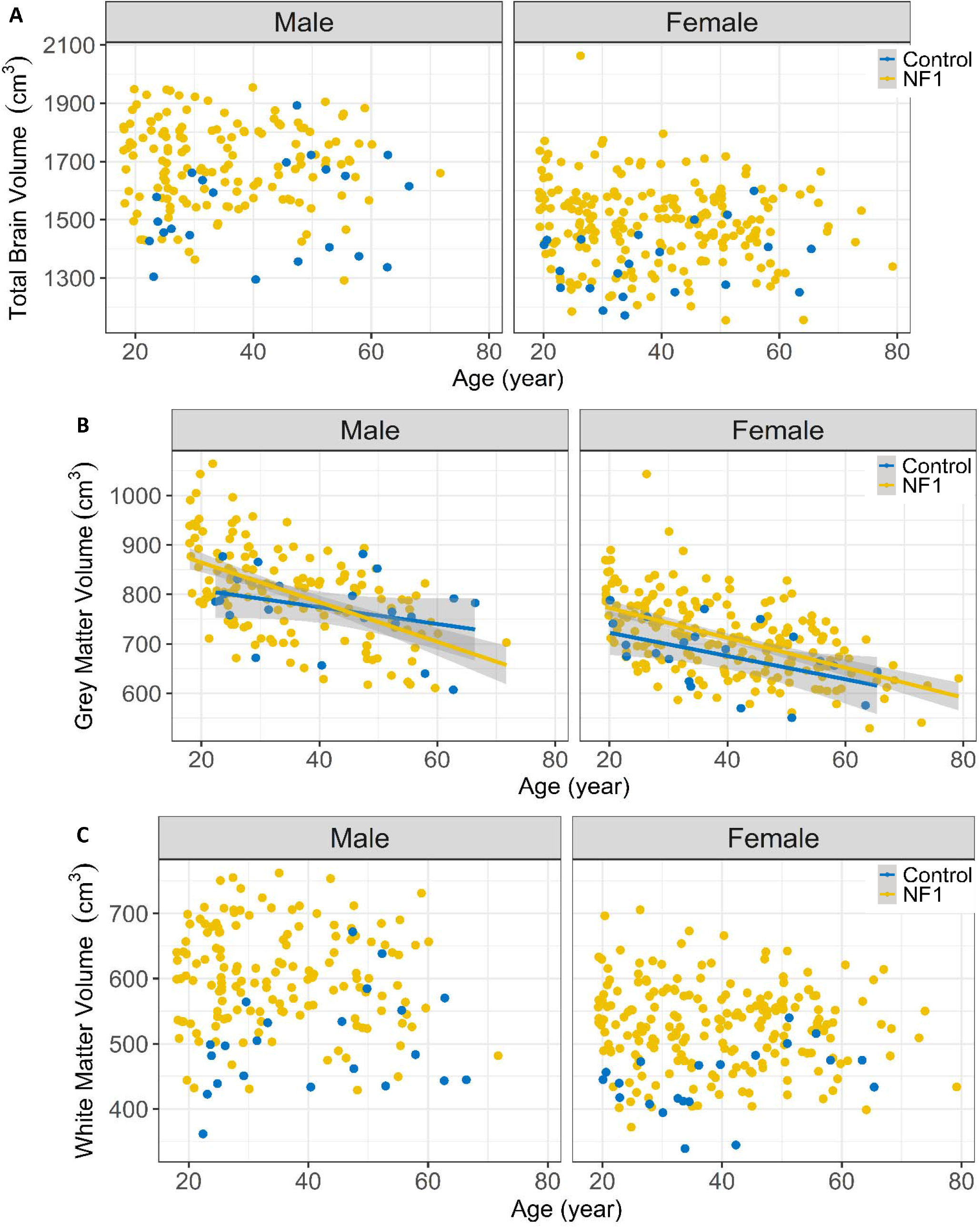
Brain volume measurements by age and sex in adults with NF1 and comparison group. (A) Total brain volume. (B) Total grey matter volume measurements. (C) Total white matter volume measurements. Linear regression lines are plotted with 1 standard error range indicated by shading. Each point represents one subject; point and line colour indicate participant group.

### Increased White Matter Volume Is Correlated with Better Psychometric Function in Adults with NF1

The average IQ of people with NF1 is reduced, and the frequency of learning disabilities and attention deficits is increased compared to unaffected individuals^3, 23, 25^. We determined whether the changes in brain volume measurements we found correlate with psychometric functional differences in adults with NF1 (Figure 5). Due to the limited sample size (N=16 to 19), we were unable to study the effect of age and sex on these correlations.

**Figure 5:**
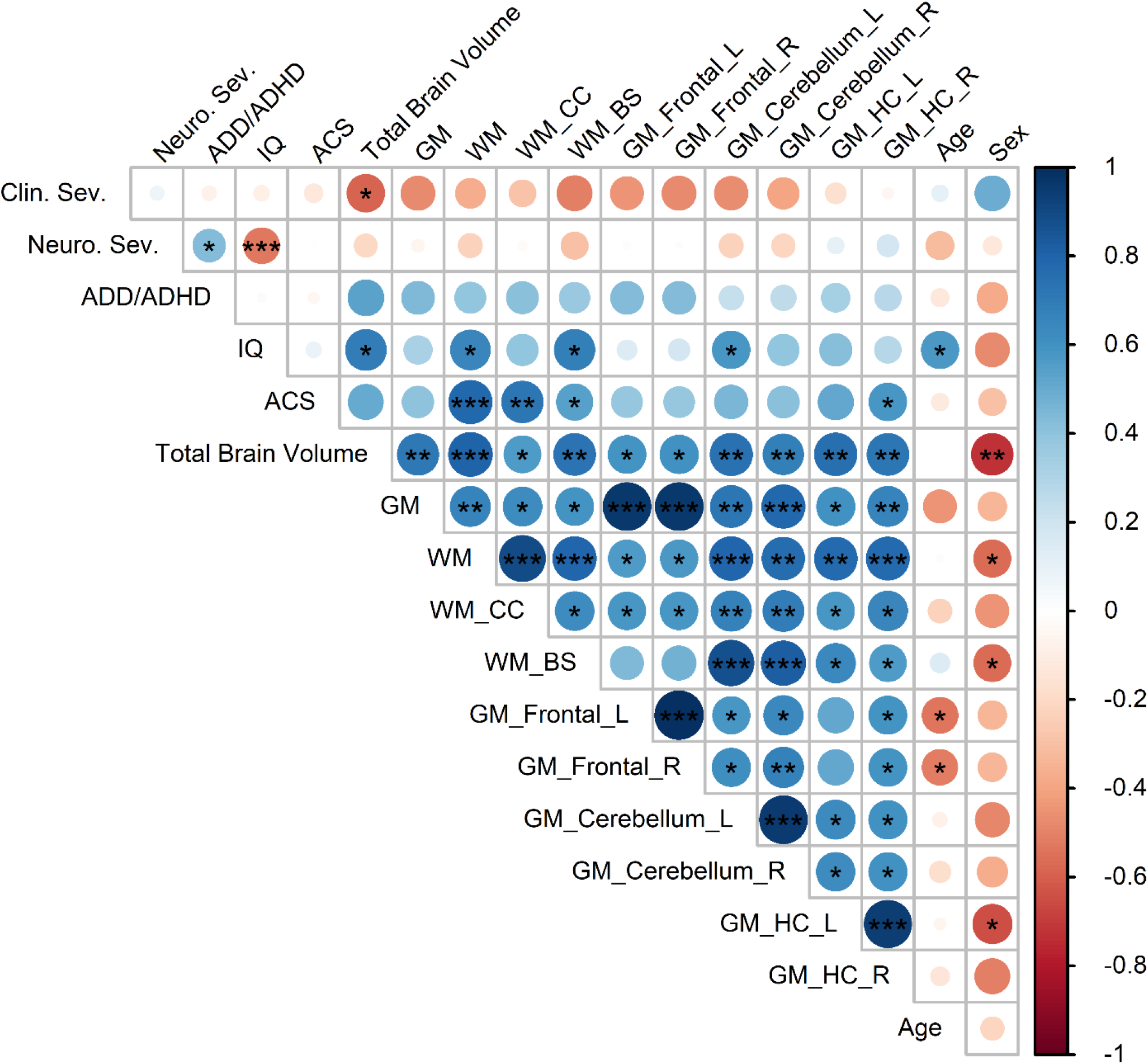
Correlation matrix of brain volume measurements with clinical and neuropsychometric assessments in adults with NF1. The color of each circle represents the degree of correlation from strongly negative (−1, red) to strongly positive (1, blue), as shown on the vertical scale on the right side of the figure. The size of the circles also indicates the strength of the correlation, with values closer to -1 or 1 larger than those that are closer to 0. Asterisks indicate FDR-adjusted statistical significance based on n = 16 – 19. * = p < 0.05, ** = p< 0.01, *** = p < 0.001. Abbreviations: Clin. Sev. = Riccardi clinical NF1 severity score, Neuro. Sev. = clinical neurological severity score, ADD/ADHD = clinical Attention deficit hyperactivity disorder severity score, IQ = intelligence quotient, ACS = Attention Comparison Score, L = left, R = right, GM = grey matter volume, WM = white matter volume, CC = corpus callosum, BS = brainstem, and HC = hippocampus without subiculum.

We found that greater total brain volume correlated with less clinically severe NF1 (*r*_*s*_ = -0.58, adjusted p = 0.024) and higher IQ (*r* = 0.69, adjusted p = 0.013). Total grey matter volume was not significantly correlated with any neuropsychometric measurements. However, regional grey matter volumes showed some correlations with psychometric function: greater cerebellum (left) grey matter volume correlated with higher IQ (*r* = 0.59, adjusted p = 0.040) and increased right HC (but not left) grey matter volume correlated with less severe attention deficiency (*r* = 0.59, adjusted p = 0.024). Higher total white matter and regional (CC and brainstem) white matter volume all correlated with better psychometric function (higher IQ and decreased attention deficit).

These results suggest that in adults of NF1, greater white matter volume might be correlated with better psychometric function. Neuropsychometric correlations with grey matter volume changes were less robust.

## DISCUSSION

The goal of this study was to investigate the effects of NF1 in adults with respect to brain volume, brain composition with respect to grey or white matter, and the correlation of brain volume differences with function. Our current findings extend our previous study, which showed that adults with NF1 exhibit enlargement of brain structures that are mainly composed of white matter, a result similar to that seen in children with NF1^22^.

NF1 is caused by mutations in the *NF1* gene, resulting in reduced ability to produce neurofibromin. This neurofibromin deficiency leads to dysregulated myelin formation in the peripheral and central nervous systems^4, 5, 41^. Previous brain volumetric studies, most of which were done in small groups of children with NF1, have usually found increased total brain volumes and total white matter volumes ^7, 8, 10, 12-14, 17-19, 21^. Our previous study, which was the first large-scale MRI investigation of unselected adults with NF1, demonstrated that the corpus callosum and mid-cerebellar peduncle (brain structures that are mainly composed of white matter), are larger than expected in NF1 patients^22^. We correlated the brain structural data to total white matter volume in a small subset of these patients and found a strong correlation between planar MRI measurements of the corpus callosum and brainstem and total white matter volume in adults with NF1^22^. These finding supported the hypothesis that the enlarged total brain volume seen in individuals with NF1 is due to an overgrowth of myelin, thus an increase in the size of white matter structures.

In the present study, we analysed volumetric MRI data in the whole brain and intracranial sub-regions of adults with NF1 and controls. We found a generalized increase in white matter volume, but we did not observe an increase in total grey matter volume or in in the volume of grey matter in the frontal, parietal, occipital, or temporal lobes. This finding differs from those in previous studies that found increased total grey matter volumes in children and young adults with NF1^7, 8, 17^. The difference in our findings may reflect a lag in brain development in younger adults with NF1, such as those studied by Karlsgodt et al. (2012)^17^, with subsequent “catch-up” of the grey matter/white matter ratio in older NF1 patients, as proposed by Moore et al. (2000)^8^. Our finding of generalized white matter enlargement in the brains of adults with NF1 is consistent with dysregulated myelin proliferation in individuals with NF1.

Brain composition changes throughout the adult lifespan, and people with NF1 have a shorter average lifespan than unaffected people^38, 42^. Thus, we were interested in the changes to white and grey matter volumes over age in adults with NF1. Our findings that age does not affect total brain volume or total white matter volume differs from observations in other patient groups, where total brain volume and white matter volume increase from early adulthood to around 45 years old then decreases afterwards^38^. The difference in our findings could be due to the small size of our comparison group (n = 43) and the paucity of adults above 45 years old in our study. We note, however, that most previously-reported studies of brain composition changes with age encompass narrow age ranges, include small sample sizes, and involve patients with various brain diseases that might affect the findings ^38^. Interestingly, we were able to show that grey matter volume decreased faster in adults with NF1 than in our comparison group.

Some studies in children with NF1 have found correlations between cognitive and behavioural impairment and CC size, total grey matter volume, or total white matter volume^8, 10, 15, 19^, although this was not found in other studies^7, 9, 14, 20, 21^. Few studies of brain morphology and neuropsychometric function have been done in adults with NF1^22^. Our morphology to neuropsychometric correlations should be interpreted cautiously because our sample size of adults with full neuropsychometric testing was quite small. As well, we were unable to study the effects of age and sex as confounding factors on our neuropsychometric correlations. In our small study, we found that increased total brain volume and white matter volume were significantly and consistently correlated with better clinical and psychometric function in adults with NF1. We did not find any correlation with total grey matter volume.

One of the main limitations of our study is the lack of 3D MRI scans for healthy adults. We used adults with NF2 or Schwannomatosis as a comparison group because a series of patient MRIs was available to us and neither of these conditions is thought to affect brain volume. We did not use published brain volumes for healthy adults as those studies included different age ranges and volumetric measurements of different regions of the brain.

Through our brain volumetric MRI analysis of the largest cohort of adults with NF1 studied to date, we showed that white matter volume is increased compared to adults with NF2 or Schwannomatosis. Thus, the increase in white matter volume previously found in children with NF1 persists into adulthood. This supports the hypothesis that NF1 causes a dysregulation of myelin production by Schwann cells in peripheral nerves and by oligodendrocytes in the central nervous system. Characterization of white matter composition and integrity using diffusion weighted imaging and studies of the relationship of the altered myelination to neuropsychometric function in adults with NF1 may help further elucidate the effects of the volumetric changes we observed on neural connectivity and function.

## Data Availability

Anonymous data are available for appropriate research purposes through V.F. Mautner, MD.

## ACKNOWLEDGEMENT

We would like to thank Sofia Granstroem who analysed the neuropsychometric data. We are grateful to the Bundesverband Neurofibromatose organization for funding this project.

## AUTHOR CONTRIBUTIONS

SW contributed to design of the study, analyzed all data, drafted and revised manuscript. JMF contributed to conception and design of the study and revised manuscript for intellectual content. PS conducted the volumetric analyses and revised manuscript for intellectual content. RB contributed to conception and design of the study, acquisition and analysis of MRI data, and revised manuscript for intellectual content. VFM contributed to conception and design of the study, recruited patients, major role in the acquisition of MRI and neuropsychometric data, and revised manuscript for intellectual content. RB and VFM contributed equally to the manuscript.

